# Quantification of microvascular lesions in the central retinal field: could it predict the severity of diabetic retinopathy?

**DOI:** 10.1101/2023.03.21.23286574

**Authors:** Jimena Fernández-Carneado, Ana Almazán-Moga, Dolores T. Ramírez-Lamelas, Cristina Cuscó, José Ignacio Alonso de la Fuente, José Carlos Pastor Jimeno, María Isabel López-Gálvez, Berta Ponsati

## Abstract

**Objective:** Diabetic retinopathy (DR) is a neurodegenerative disease characterized by the presence of microcirculatory lesions. Among them, microaneurysms (MAs) are the first observable hallmark of early ophthalmological changes. The present work aims at studying whether the quantification of MA, haemorrhages (Hmas) and hard exudates (HEs) in the central retinal field (CRF) could have a predictive value on DR severity.

**Research Design and Methods:** Retinal lesions (MAs, Hmas and HEs) were quantified in the CRF of 160 retinographies from diabetic patients from the IOBA’s reading center, previously classified by two expert readers with the 3 fields-Joslin system. Samples included different disease severity levels and excluded proliferating forms: no DR (n=30), mild non-proliferative (n=30), moderate (n=50) and severe (n=50).

**Results:** Quantification of MAs, Hmas, and HEs revealed an increase trend of these lesions as DR severity progresses. Differences between severity levels were statistically significant, suggesting that the analysis of the CRF provides valuable information on severity level and could be used as a valuable tool to assess DR grading in the clinical practice.

**Conclusions:** Even though further validation is needed, the counting of microvascular lesions in the central retinal field can be proposed as a rapid screening system to classify DR patients with different stages of severity according to the international classification.

## INTRODUCTION

A total of 537 million people worldwide were estimated in 2021 to have Diabetes Mellitus (DM), representing 10.5% of the global adult population (20–79 years). This number is expected to increase to 643 million (11.3%) in 2030 and up to 783 million (12.2%) in 2045.^1^

A very frequent complication is Diabetic Retinopathy (DR), classically considered a microcirculatory disease, but currently defined also as a neuropathy.^2^ DR has become the leading cause of blindness in working-age adults (20-65 years) in the most developed countries. In fact, approximately 30% of people with DM develop some degree of DR.^3^ Metabolic control, through the monitoring of blood glucose and glycated hemoglobin (HbA1c) levels, has substantially contributed to arrest DR progression, however, the high prevalence of visual impairment and blindness has increased since 1990, mainly due to type II diabetes, as a consequence of a growing and ageing diabetic population.^4^

DR falls into two categories depending on the presence of neovascularization (NV) in the retina, being classified as non-proliferative (NPDR) when there is no evidence of abnormal blood vessel growth, or proliferative (PDR), when neovascularization occurs.^5^ NPDR is further characterized by the presence of different types of retinal vascular lesions, such as microaneurysms (MAs), haemorrhages (Hmas), hard exudates (HEs), cotton-wool spots (CWS), intraretinal microvascular abnormalities (IRMA) and venous beading (BV), among others.^6^ The combination of these retinal lesions and their number define the severity of a NPDR patient. Although a wide range of lesions can appear in a diabetic retina, the earliest clinical sign of DR are usually MAs, which are small dilations of the capillaries as a consequence of the loss of pericytes, apoptosis of endothelial cells accompanied of thickening of the basement membrane and capillary occlusion.^7^ MAs are clinically identified as small red dots, with increased permeability, located in the small retinal vessels.^8^ They may bleed and result in Hmas, which can also appear independently, in different shapes and sizes depending on their location. MAs are known to be highly dynamic lesions, meaning that, over the course of the disease, some of them may disappear, as a consequence of spontaneous occlusion and progressive remodelling of retinal vasculature.^9^ Vascular damage can also lead to the leakage of fluid and lipoproteins into the outer plexiform layer, forming intraretinal edema and the so-called HEs. They are irregularly shaped yellow-white spots that can coalesce with each other forming streaks or clusters most of them centered by microvascular leaking structures.^6^

As mentioned, the appearance of MAs is considered a hallmark of early changes in the retina, so they are considered as the first pathological clinical sign for ophthalmoscopic DR diagnosis.^10,11^ MAs analysis has been the focus of interest because different studies have demonstrated that the presence of MAs closely centred on the macula may be associated with complications that cause visual impairment by worsening DR or Diabetic Macular Edema (DME) and onset of PDR.^12^ In fact, several studies have demonstrated that total number of MAs as well as MAs turnover rate (MAT) (i.e. MA formation rate plus MA disappearance rate) in the central area of the retina are predictors of disease progression suggesting that even a single MA may have a predictive value in DR progression.^13,14^ In this sense, the Wisconsin Epidemiologic Study of Diabetic Retinopathy in 1989 was the first to demonstrate that the number of MAs at baseline plays an important role as predictor of progression of DR.^15^ Later, Klein et al. established in 4- and 10-year follow-up studies of mild NPDR patients with only MAs at baseline the relationship between higher MAs number and the progression of DR to moderate NPDR which is a clinically relevant severity level from which the risk of subsequent development of PDR or Clinically Significant Macular Edema (CSME) is appreciable.^16^ Kohner et al. also confirmed that MAs number has a highly predictive value for worsening DR in a 12-year follow-up study from same population.^10^

DR is currently classified at daily practice in four severity levels: mild, moderate, severe NPDR and PDR, following the International Clinical Diabetic Retinopathy (ICDR) Disease Severity Level.^17^ Fundus examination of the eye by an expert ophthalmologist, without capturing images, is still nowadays the most common practice of the ophthalmological examination to diagnose DR and classify its severity degree.^3,18^

Outside the daily practice, in clinical trials (CT), the stages of DR have been further detailed and classified by ETDRS. To do this, 7-standard field CFP stereoscopic images are acquired to detect the presence of specific lesions in one or more retinal fields and its relative intensity compared to standard CFP images. The combination of both parameters leads to a numerical code that represents the severity level and its association with the occurring retinal lesions^19^ (**Table 1**).

**Table 1.**
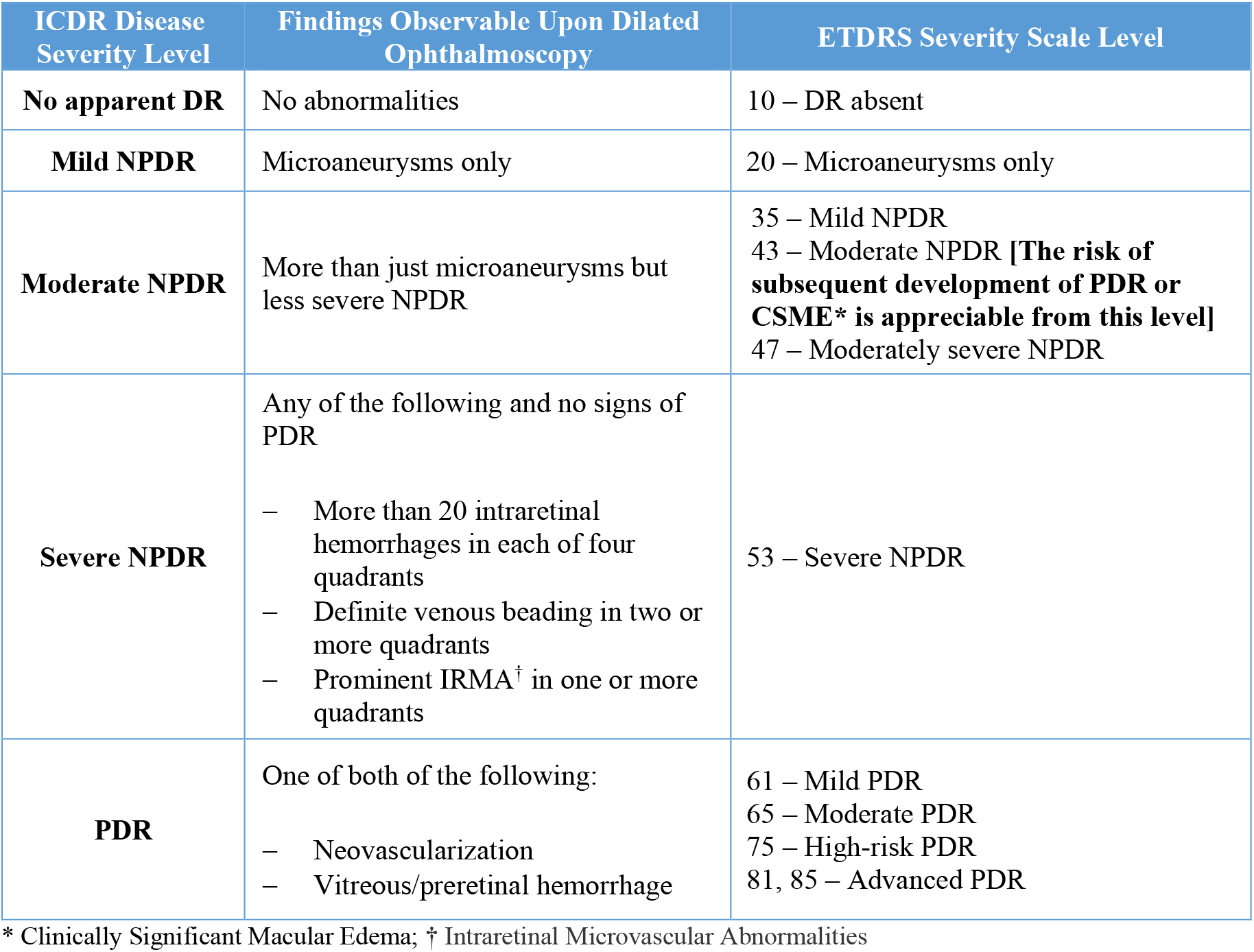
Comparison between International Clinical Diabetic Retinopathy (ICDR) Disease Severity Level and Early Treatment Diabetic Retinopathy Study (ETDRS) Severity Scale level (adapted from Wilkinson et al., Ophthalmology 2003;110:1677–1682)

| ICDR Disease Severity Level | Findings Observable Upon Dilated Ophthalmoscopy | ETDRS Severity Scale Level |
| --- | --- | --- |
| **No apparent DR** | No abnormalities | 10 – DR absent |
| **Mild NPDR** | Microaneurysms only | 20 – Microaneurysms only |
| **Moderate NPDR** | More than just microaneurysms but less severe NPDR | 35 – Mild NPDR<br>43 – Moderate NPDR [**The risk of subsequent development of PDR or CSME\* is appreciable from this level**]<br>47 – Moderately severe NPDR |
| **Severe NPDR** | Any of the following and no signs of PDR<br>– More than 20 intraretinal hemorrhages in each of four quadrants<br>– Definite venous beading in two or more quadrants<br>– Prominent IRMA<sup>†</sup> in one or more quadrants | 53 – Severe NPDR |
| **PDR** | One of both of the following:<br>– Neovascularization<br>– Vitreous/preretinal hemorrhage | 61 – Mild PDR<br>65 – Moderate PDR<br>75 – High-risk PDR<br>81, 85 – Advanced PDR |

\* Clinically Significant Macular Edema; † Intraretinal Microvascular Abnormalities

The ETDRS grading protocol is a semi-quantitative method used to subclassify the severity degree of the DR patients enrolling a CT because it is assumed to provide more accuracy. It allows the evaluation of additional changes within the same ICDR disease severity level.

Although ETDRS is the gold standard and the current metrics for regulatory assessments, it has not been implemented in the ophthalmological practice due to practical reasons. Firstly, the need for the adequate technical equipment and skilled personnel trained to correctly acquire the 7-field CFP images of the retina, which includes the challenges of capturing appropriate images of peripheral fields. Secondly, the need for experienced ophthalmologists able to differentiate between sequential severity levels, e.g., patients with moderate DR (ETDRS 35, ETDRS 43 or ETDRS 47 level), and, finally, the requirement for patient cooperation.^20^

Since DR remains asymptomatic until it has significantly advanced affecting the vision in the central area, screening to detect it during the early stages and new indicators for assessing the progression or amelioration of the disease are needed.^21,22^

Bursell et al. validated in 2001 a new protocol to simplify the examination and diagnosis of DR severity, known as the Joslin Vision Network protocol (JVN).^23^ Instead of 7 fields, this system uses 3 non mydriatic (NM) 45°-field stereoscopic colour images (named NM 1, NM-2 and NM-3). When compared to the use of the 7 standard CFP in the screening of DR, the sensitivity and specificity for detecting referable levels of DR were 82% and 92% for the 3-fields (NM-1, NM-2 and NM-3) and 71% and 96%, respectively, for the central field NM-1 alone. This 3-field system was found to be as effective for DR screening and severity classification (in Mild, Moderate, Severe and PDR levels) as the standard 7-fields 30-35-mm fundus photographs, so it was implemented due its advantage in terms of training, time and cost.^20^

Further attempts to reduce the number of images needed for determining the presence of DR and defining the severity level have been done. In this sense, the approach of two 45° digital CFP is used in the first autonomous artificial intelligence (AI)-based device for automatic diagnosis of RD with FDA approval for its use in clinical practice.^24^ Other studies have evaluated the effectiveness of a single field macula-centred 45° digital CFP. These studies demonstrated a sensitivity and specificity for DR screening between 70-95% and have been translated into clinical benefit with the approval of some AI-based DR detection tools.^25^ However, although recording only the central field simplifies the procedures, a single image of the retina is not considered representative enough for the definition of the severity level in clinical practice.

## RESEARCH DESIGN AND METHODS

### Hypothesis

Based on the fact that DR microvascular changes can be diagnosed only by detecting the presence of at least 1 MA in a single CFP image, we hypothesize that findings in the central retinal area could predict with sufficient accuracy the severity levels estimated by the JVN protocol. This fact has the pathophysiological basis that the central area is the most susceptible to metabolic alterations of DM.^26,27^

Our objective was to study the correlation between the number of microvascular lesions in a central CFP image and the DR severity level in NPDR patients, previously classified by two expert readers following the 3-field CFP JVN system.

The hypothesis of the present work is that if the correlation is confirmed, the quantification of MAs, Hmas, both or HEs in the central CFP field could be considered in clinical practice as a first metrics for a fast evaluation of the DR severity degree.

### Study design and Study population

The CFP images were provided by the *Instituto de Oftalmobiología Aplicada* (IOBA) of the University of Valladolid (IOBA-UVA) reading centre from their image data set. IOBA-UVA reading centre holds a CFP database, from the DR blindness prevention program of the Junta de Castilla y León (Regional Government), with images captured and read by certified personnel using the JVN system. Images were captured using a Topcon TRC-NW400 automatic retinal camera (Topcon Medical Systems, Inc., Oakland, NJ, USA) at a 45-degree field. The photograph’s acquisition protocol was performed under mydriasis with tropicamide. This is a retrospective study in which all subjects signed their informed consent for the transferring of their data and images for teaching and research purposes. Moreover, this research was carried out under full compliance of the General Data Protection Regulation (GDPR).

A total of 160 anonymized images of type 1 and 2 DM patients were analysed. Patients with PDR were excluded. The manual quantification of retinal lesions (MAs, Hmas and HEs) was conducted by two independent, experienced and certified ophthalmology graders evaluators from IOBA-UVA reading centre and it was performed in the CFP NM-1. Four groups of images were established: No RD (n=30), mild NPDR (n=30), moderate NPDR (n=50) and severe NPDR (n=50). MAs were defined as red lesions (RLs) saccular dilations with well-defined and identifiable borders, and Hmas were defined as RLs of variable morphology (punctate, flame, irregular edges). HEs were identified following a protocol of morphological characteristics.^28^ **Figure 1** shows an example of a CFP image illustrating the presence of MAs, Hmas and HEs.

**Figure 1.**
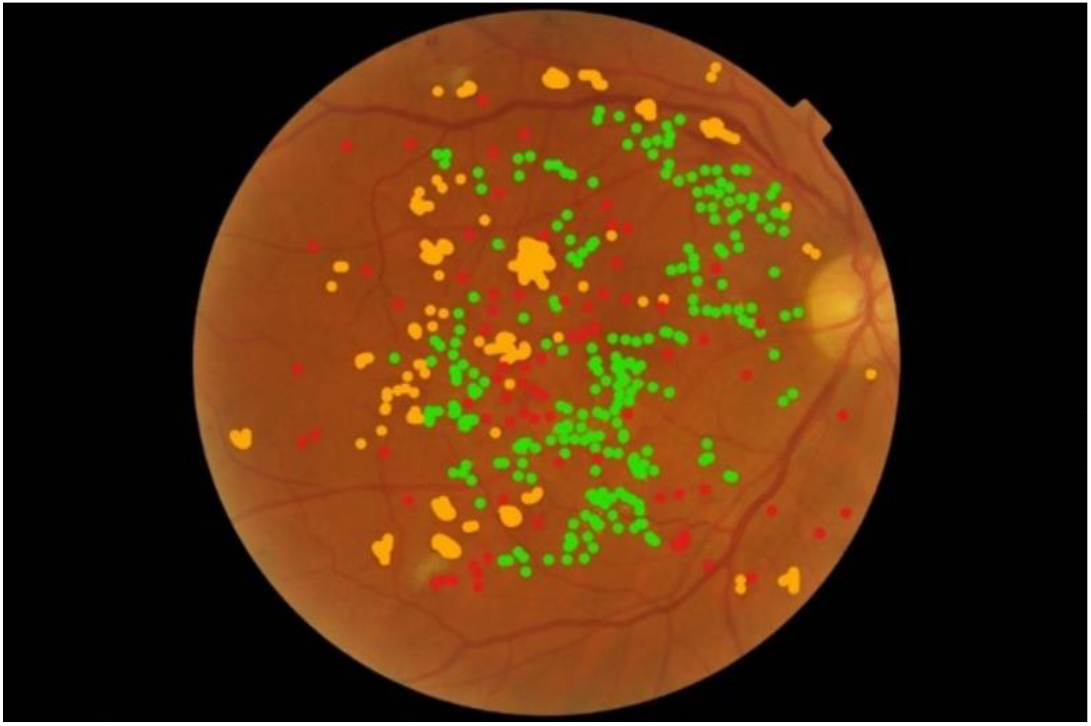
Example of a CFP image with MAs marked in red, Hmas in orange and HEs areas in green

### Statistical analyses

Statistical analysis was performed by using Graphpad Prism 8.0.1. Descriptive statistics (including number of values, mean, standard deviation (SD) and standard error of the mean (SEM)) of each microvascular lesion for each DR degree level were calculated for all parameters. The intergroup (unpaired T-test) differences were performed (two-sided, α=0.05)

## RESULTS

The number of MAs, Hmas, RLs and HEs quantified in the central NM-1 field were plotted separately against each DR severity level (no DR, mild, moderate and severe NPDR), as shown in **Figure 2**.

**Figure 2.**
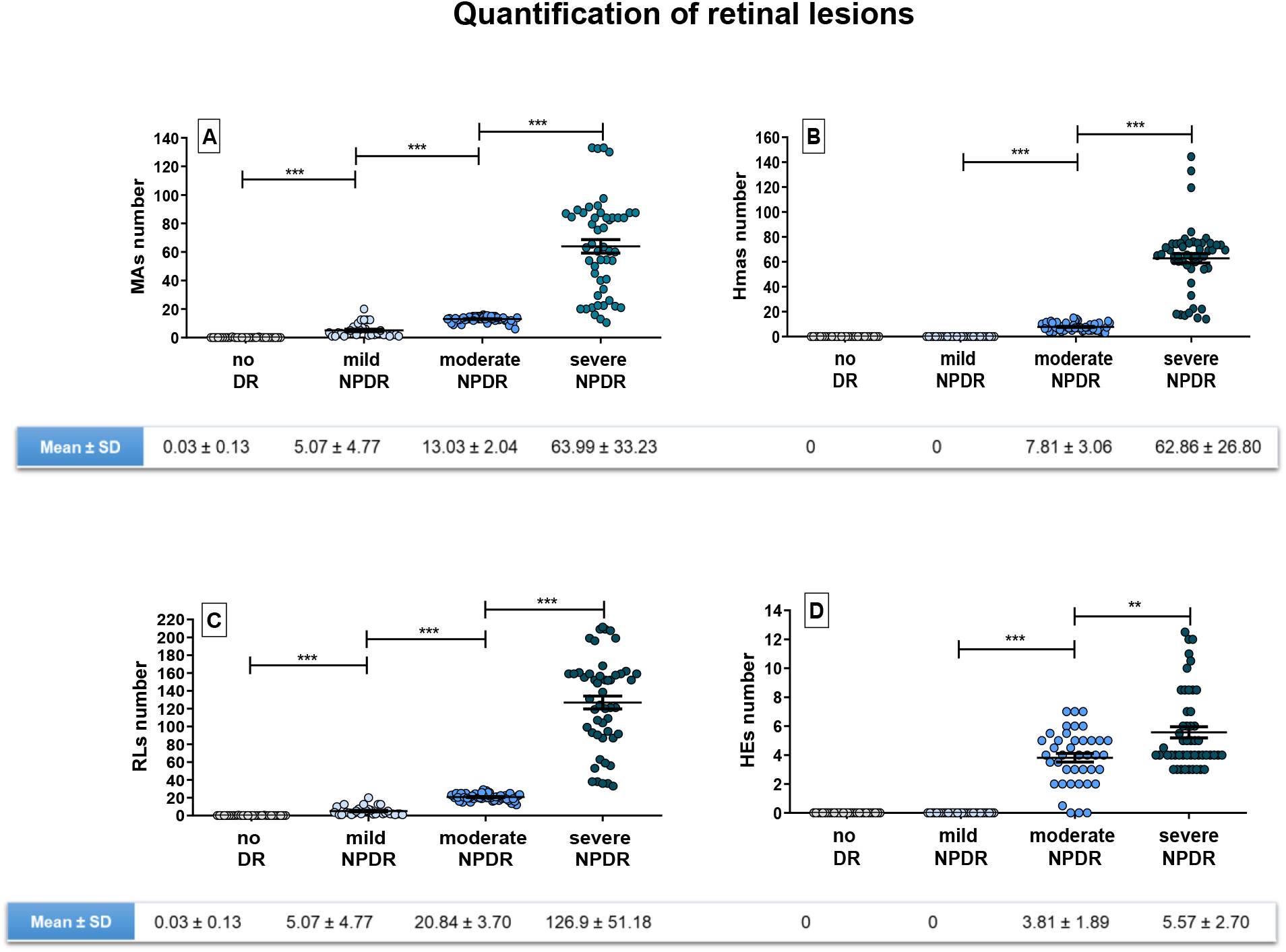
Quantification of a specific type of retinal lesions in a single macula-centered NM-1 field, plotted against severity level (no DR, mild NPDR, moderate NPDR and severe NPDR. A) Number of MAs quantified; B) Number of Hmas quantified; C) Number of RLs quantified, defined by the sum of MAs and Hmas; D) Number of HEs quantified. Below each graph the mean ± SD is shown. Level of significance: *** p< 0.0001 and ** p<0.001.

According to **Figure 2**, the number of MAs quantified in the NM-1 central image of patients with no DR was negligible, while mean MAs number in mild, moderate and severe NPDR patients was 5.07 ± 4.77, 13.03 ± 2.04 and 63.99 ± 33.23, respectively. According to these results, the mean MAs number significantly increased with the DR severity stage (no DR < mild DR < moderate DR < severe DR; p<0.0001).

Regarding Hmas, they were detected in moderate and severe NPDR patients and quantified as 7.81 ± 3.06 for moderate patients and 62.86 ± 26.80 for severe cases.

RLs, representing the combination of the mean values of both MAs and Hmas, showed the same trend as MAs quantification alone, but with higher numbers due to the influence of Hmas (0.03 ± 0.13 for no DR; 5.07 ± 4.77 for mild; 20.84 ± 3.7 for moderate and 126.9 ± 51.18 for severe NPDR).

Finally, as expected, no HEs were identified in patients without or mild NPDR, while the values increased to 3.81 ± 1.89 for moderate cases and 5.57 ± 2.70 for severe ones. The difference on the quantification of retinal lesions between severity levels was statistically significant in all cases.

Besides considering the lesions separately, we also analyzed the quantifications by combining all types of lesions (MAs, Hmas, HEs) in the same plot, as shown in **Figure 3**.

**Figure 3.**
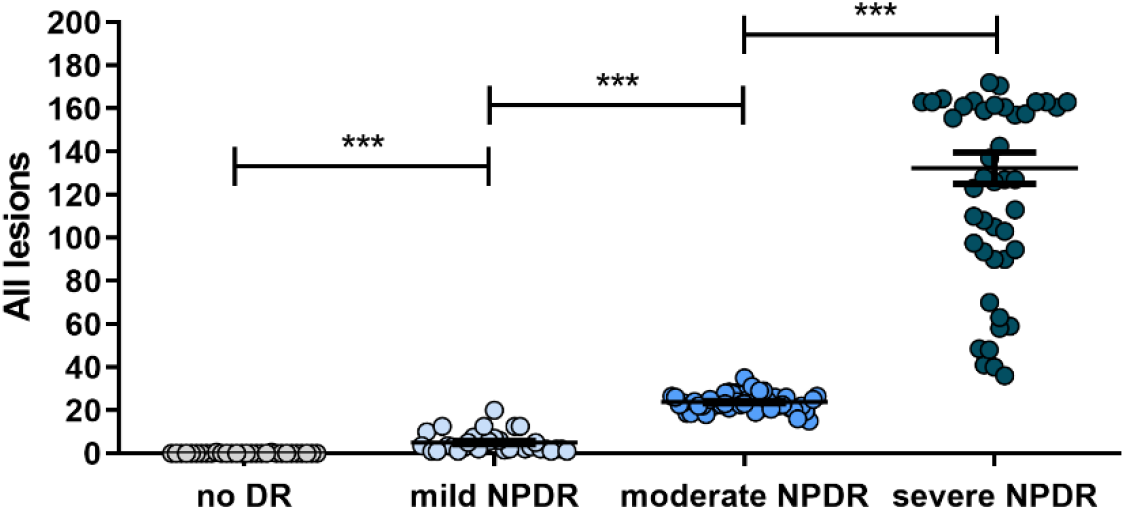
Quantification of all lesions in a single macula-centered NM-1 field, plotted against severity level (no DR, mild NPDR, moderate NPDR and severe NPDR. Level of significance: *** p< 0.0001.

The results obtained as the global microvascular picture (**Figure 3**), considering the lesions altogether, indicate that their appearance in a single NM-1 central field significantly increases with DR severity.

## CONCLUSIONS

The results of the quantification of MAs, Hmas, RLs and HEs revealed an increase trend of these vascular lesions with the DR severity level, as expected. Noteworthy, the differences between the number of lesions counted in the NM-1 field among severity degrees were statistically significant, suggesting that the detailed analysis of this single field could provide valuable information on the severity level. This leads to the consideration that, as MAs are the first observable hallmark of DR and their presence is differential across the severity stages, they could be proposed as a potential marker for assessing DR level.^29^ Nevertheless, these results require further validation.

Interestingly, the quantitative ultra-widefield (UWF) fluorescein angiographic metrics reported by Ehlers et al. also concluded that MAs quantification, panretinal in that case, is also associated with DR severity.^30^ Fluorescein angiography is valuable for quantifying MAs and differentiating them from Hmas, but its implementation for automated follow up in the ophthalmologic practice is not foreseeable due to its invasiveness.^31^

Other publications are consistent with our results, as the one by Xu et al. on the automated quantification of Hmas in CFP images captured with a non-mydriatic camera, based on an intelligent system.^32^ Their study highlighted the correlation between the number of haemorrhagic lesions and the DR severity in both mild and moderate NPDR patients, supporting the current results.

Other studies have also drawn the attention to microvascular abnormalities, such as MAs, as a potential and unbiased tool to quantitatively monitor DR progression. In this case, authors report the advantages of Optical Coherence Tomography Angiography (OCTA) as a safe alternative to examine the effect of DR on individual retinal layers and diagnose DR even before it is clinically detectable on ophthalmoscopic fundus examination.^11^

Although it is indeed an appealing technique, we emphasize in finding a simplified approach that enables the evaluation of microvascular abnormalities only in the central field of CFP images since the appearance of microvascular lesions close to the macula is recognised in NPDR.^33^

It is interesting the HEs data dispersion (**Figure 2**) in comparison with the lesions of vascular origin in the stage of moderate NPDR which may be related to the difficulty of identifying these lesions, which usually form plaques, in an isolate way. In fact, it has recently been proposed to measure the area they occupy instead their number.^34^

While the development of new AI or deep learning algorithms for automatic image analysis aiming to differentiate MAs and Hmas is on-going,^25^ their manual quantification is still challenging. In fact, the distinction between both lesions in CFP images is complicated and even impossible in many cases. A plausible solution to this issue, could be reporting MAs and Hmas together as Red Lesions (RLs), becoming potential metrical approach for disease characterization.

Although this work presents the potential of the quantification of specific retinal lesions in a single field as an approach to assess the level of DR severity, it has also limitations due to the relevance of peripheral fields in many cases of DR. Li et al. demonstrate that the typical clinical signs of DR, including MA, IRMA, capillary non-perfusion areas and NV, are commonly distributed in the inferior nasal mid-peripheral areas besides the posterior pole, assessed by fluorescein angiography and CFP.^35^ This means that the microvasculature along retinal periphery needs to be adequate considered in order to have a more accurate DR evaluation. Fortunately, retinal photography and imaging has progressed substantially over the past decade, so new commercially available UWF systems have been developed, allowing imaging 200º of the retina in a single image, contrasting the 30º or 45º achieved in the ETDRS standard protocol or the JVN NM-I system.^36^

In this line, Silva et al. report that UWF imaging is able to reveal substantially more diabetic retinal vascular pathology, even without the use of fluorescein angiography. In fact, the authors show that one third of Hmas/MAs, IRMA and NV are located predominantly outside the 7 fields ETDRS, suggesting that the severity level can be misleading.^37^ In another study, Silva et al. highlighted the benefit of objective quantification for DR assessment for a more precise DR scoring system. The authors evaluate retinal lesions in UWF images based on subjective (performed by 2 masked graders) and quantitative assessments (lesion frequencies and surface area) and reveals that 22% of the identified lesions are distributed outside the ETDRS fields.^38^ In the same way, Sadda et al. show that patients that have undergone PDR in 4 years are prone to show more retinal lesions at the peripheral level.^39^

Although the present work lacks the possibility to evaluate the periphery of the retina, the acquisition of a single-field central CFP could be of high clinical value for a fast assessment of DR based on quantifiable variables. In fact, the use of numerical metrics over semi-quantitative or qualitative parameters could potentially allow the detection of observable improvements within the same level of severity, being useful for evaluating the effectiveness of treatments, for instance. Moreover, since the evaluation of only one field gives differential information depending on the DR severity level, its acquisition seems interesting for following the patients in the eye care routine, giving breath to the current cost- and resource-constrained healthcare systems. Finally, this affordable method shows the advantage that the vast majority of ophthalmological settings have standard non-mydriatic fundus cameras,^40^ while not all of them are equipped with UWF imaging.

In this sense, a standard simplified protocol for the daily practice should be agreed and deployed in the clinical health system based on CFP. The development of such clinical guideline, however, would be challenging due to different factors, starting from the choice of mydriatic (affording higher resolution images) versus non mydriatic cameras (a better option to avoid increase of intraocular pressure), the selection of the most suitable retinal field, and according to which system (7-field ETDRS, 3 NM JVN fields, etc.), among others.

This leads to the conclusion of Solomon et al. and Simó et al.^21,22^ on the need for international ophthalmology leaders to systematically address how to quantify DR disease progression and to define the best techniques and protocols. In this sense, our goal is to make our results available to experts for their consideration as exploitable and quantifiable variables for DR detection and decision making, based on microvascular counting of MAs and Hmas. Of course, these results require validation with an external sample before approving them as a rapid screening system, which would be very accessible to the daily clinic.

As far as we are concerned, the present work is the first study that shows a statistically significant difference in the number of MAs, Hmas and RLs between DR severity levels quantified only in the central CFP. Although it requires further validation, the quantification done only in the central field of CFP, shows a robust correlation between the number of the aforementioned lesions and DR severity level.

Furthermore, the potential of MAs and Hmas as quantifiable variables in fundus images for the disease severity level could proposed as clinically relevant efficacy endpoints for clinical trials.

In conclusion, our results could open up new tools to be implemented in the ophthalmological daily practice for recording the pathway of the disease and also for evaluating new therapeutic avenues for treating DR.

## Data Availability

The Color Fundus Photography images were provided by the Instituto de Oftalmobiologia Aplicada (IOBA) of the University of Valladolid (IOBA-UVA) reading centre (Valladolid, Spain) from their image data set. IOBA-UVA reading centre holds a CFP database, from the DR blindness prevention program of the Junta de Castilla y Leon (Regional Government), with images captured and read by certified personnel using the JVN system.

## Ethical Approval

The Ethics Committee (CEIm) from Á*rea de Salud Valladolid Este* gave ethical approval for this research project.

## Author Contributions

J.F.C, A.A.M, D.R.L, C.C and B.P. contributed to the general coordination, design and conceptualization of the study, participated in the interpretation of the results and the preparation and review of the manuscript.

J.I.A, J.C.P. and M.I.L. contributed to the research data, image quantification, and the writing and review of the manuscript.

All authors approved the final version of the manuscript.

## Guarantor Statement

J.C.P. and M.I.L. are the guarantors of this work and, as such, had full access to all the data in the study and take responsibility for the integrity of the data and the accuracy of the data analysis.

